# Functional Dysconnectivity of White Matter Networks is Associated with Clinical Impairment in Autism Spectrum Disorder

**DOI:** 10.64898/2026.03.08.26347903

**Authors:** Si-Jing Wu, Miao-Ting Huang, Dan-Wei Huang, Ze-Qiang Linli, Shui-Xia Guo

## Abstract

**Background:** Structural white matter (WM) alterations are recognized in Autism Spectrum Disorder (ASD), yet the functional connectivity (FC) of WM networks and its clinical significance remain largely under-explored.

**Methods:** This study aimed to investigate aberrant FC patterns within intra-WM (WM-WM) and WM-gray matter (WM-GM) networks in a large ASD cohort. Resting-state fMRI data from 272 ASD individuals and 368 typical controls (TC) from the ABIDE-II dataset were analyzed. We constructed WM-WM and WM-GM FC networks using Pearson correlations between atlas-defined regions, applied ComBat harmonization, and employed Network-Based Statistics (NBS) to identify group differences. Associations with clinical symptoms were assessed using Social Responsiveness Scale (SRS) scores, and a CatBoost algorithm was used for diagnostic classification based on connectivity features.

**Results:** NBS analyses revealed significantly increased connectivity in ASD for 116 WM-WM pairs and 58 WM-GM pairs (P<0.05, FWER-corrected). Critically, the strength of these aberrant WM-WM functional connections exhibited a significant negative correlation with SRS total scores (r = -0.22, P < 0.001), whereas WM-GM connectivity showed no such significant association. The hybrid CatBoost classifier, integrating both WM-WM and WM-GM features, achieved moderate diagnostic discrimination (AUC = 0.669 ± 0.040).

**Conclusion:** These results offer novel insights into the aberrant functional architecture of WM-related networks in ASD, particularly linking intra-WM dysconnectivity to symptom severity, thereby enhancing our understanding of the neural substrates underlying social-communicative deficits.

## 1. Introduction

Autism Spectrum Disorder (ASD) is a neurodevelopmental condition characterized by core behavioral deficits in social interaction, verbal communication, and repetitive behaviors. With its global prevalence rising steadily, ASD poses a significant public health challenge. Resting-state functional magnetic resonance imaging (rs-fMRI) studies have provided valuable insights into the effects of large-scale brain functional connectivity (FC) by measuring blood oxygenation-level dependent (BOLD) fluctuations in fMRI signal (Cai et al., 2024; Gore et al., 2003; Biswal et al.,1995). Recent studies have identified significant FC alterations in ASD. For example, ASD is associated with reduced Regional Homogeneity (ReHo) in the superior temporal gyrus and the Middle Occipital Gyrus (Yue et al., 2023); increased FC between the occipital lobe and precuneus; and decreased FC between the Angular Gyrus (ANG), Middle Temporal Gyrus (MTG), and the parietal lobe and precuneus (Randeniya et al., 2023; Xu et al., 2019). Notably, the majority of rs-fMRI research has exclusively focused on gray matter (GM) networks, while white matter (WM), a critical neural substrate for transmitting neural signals between GM regions (GM-GM FC), has been largely ignored. This gap can be partly attributed to the systematic removal of WM BOLD signals as “noise” during preprocessing.

However, increasing evidence indicates the neurophysiological relevance of WM BOLD signals and justifies their inclusion in functional connectivity research. Key findings include: (1) WM BOLD signals are modulated by neural activity and exhibit meaningful spatiotemporal dynamics (Ding et al., 2018; Schilling et al., 2023); (2) WM-FC corresponds with diffusion tensor imaging-based tractography and correlates with GM networks, supporting its structural basis (Peer et al., 2017); (3) WM functional networks are reproducible and exhibit genetic underpinnings (Li et al., 2021); and (4) WM dysfunction has been linked to multiple neuropsychiatric conditions, underscoring its clinical relevance (Ji et al., 2023; Chen et al., 2023b; Cui et al., 2021).

Notably, in the context of ASD, structural MRI and diffusion tensor imaging (DTI) studies have consistently revealed WM microstructural alterations, including reduced volume and impaired axonal integrity in circuits underlying social cognition (Gao et al., 2021; Andrews et al., 2021; Koshiyama et al., 2020). These early developmental WM abnormalities are hypothesized to be either primary etiological factors or critical contributors to ASD pathogenesis. Despite converging evidence from structural WM alterations, the FC alterations in ASD related to WM remain elusive.

Accordingly, this study capitalizes on the large-scale ABIDE-II dataset to conduct a comprehensive investigation into the functional organization of WM networks in ASD. By constructing both intra-WM and WM-GM networks, we aimed to answer three central questions: First, do individuals with ASD exhibit distinct patterns of intra-WM and WM-GM functional connectivity compared to typically developing controls? Second, are these network-level aberrations associated with the clinical severity of autistic traits? Third, do these WM connectivity features hold diagnostic potential for classification? Collectively, this work aims to refine the neurobiological model of ASD by integrating the functional dynamics of the brain’s white matter.

## 2. Materials and Methods

### 2.1 Participants

This study utilized rs-fMRI and phenotypic data obtained from the ABIDE II database. Following standardized quality control protocols, the final analytical sample consisted of 640 participants matched for gender distribution, comprising 272 individuals with ASD and 368 typical controls (TC). Core autism symptomatology was assessed using the Social Responsiveness Scale (SRS). The total raw score was utilized as a continuous measure of global social functioning impairment, with higher values indicating greater deficits. Additionally, the SRS also evaluates three principal diagnostic constructs: Social Communication and Interaction (COMM), Awareness and Responsiveness (SOCIAL), and Restricted/Repetitive Behavioral Patterns (RRB). These domains respectively correspond to the symptomatic manifestations of Autism Spectrum Disorder in the areas of communication capacity, social functioning, and stereotyped behaviors. Table 1 presents the demographic characteristics of the ASD and TC groups.

**Table 1.** Demographic and clinical characteristics of participants.

| **Variable** | **ASD (n = 272)** | **TC (n = 368)** | **P-value** |
| --- | --- | --- | --- |
| Age (years) | 15.75 (10.48) | 15.44 (NA) | 0.702 |
| Sex, n (%) | Male: 233 (85.7%)<br>Female: 39 (14.3%) | Male: 269 (73.1%)<br>Female: 99 (26.9%) | <0.001 |
| Mean Framewise Displacement (mm) | 0.11 (0.06) | 0.09 (0.05) | <0.001 |

Note: Values are presented as mean (standard deviation) unless otherwise indicated. P-values were obtained using independent samples t-tests for continuous variables and chi-square tests for categorical variables.

### 2.2 Data Processing and Quality Control

All neuroimaging data were preprocessed and subjected to rigorous quality control pipelines implemented in MATLAB using SPM12 (Statistical Parametric Mapping; http://www.fil.ion.ucl.ac.uk/spm/software/spm12) and DPARSF (Data Processing Assistant for Resting-State fMRI; http://rfmri.org/DPARSF). The preprocessing of the T1 images included segmenting WM, GM, and cerebrospinal fluid in each subject’s native anatomical space, registering each T1 segmentation image to the functional space of each subject, and constructing individual-level WM or GM masks. In this initial tissue segmentation stage, a probability threshold of 0.5 was used to generate preliminary tissue masks, aiming to maximize the retention of potential tissue-specific signals. The preprocessing of the fMRI data included the following key steps: (1) Removal of the first five time points. (2) Slice timing correction and head motion correction. (3) Co-registration of the functional images with the structural images. (4) Removal of linear trends. (5) Nuisance regression using 24 motion parameters and mean cerebrospinal fluid signal (WM and global signals were retained for WM-FC analysis); (6) Time scrubbing using motion “spikes” (FD>2mm) as individual suppression factors. (7) Bandpass filtering was set to 0.01–0.10 Hz for GM and 0.01–0.15 Hz for WM to capture relevant spectral characteristics in WM regions; (8) Spatial smoothing applied separately within WM and GM masks to prevent mixing of signals (FWHM = 4 mm); (9) Normalization of functional images to Montreal Neurological Institute (MNI) space using Diffeomorphic Anatomical Registration Through Exponentiated Lie Algebra (DARTEL) and resampling to 3×3×3 mm^3^ voxels. Finally, signal extraction based on the atlas ROIs was performed on these MNI-normalized functional images.

A manual quality control procedure was performed after preprocessing. The following criteria were used to assess data quality: (1) successful generation of all preprocessed results; (2) mean FD below 0.2 mm; (3) visual inspection confirming satisfactory spatial normalization; and (4) each site included more than 30 participants. Subjects with incomplete data at any site were excluded prior to harmonization. The final analysis included 640 participants (272 ASD and 368 TC).

### 2.3 Construction of Functional Networks and Harmonization

To generate both WM-GM and WM-WM resting-state functional connectivity (rsFC) matrices for each subject, Pearson’s correlation coefficients were computed between regional time courses of fMRI signals from predefined atlas-defined WM tract ROIs and GM parcels, as well as between WM tract ROIs themselves. These coefficients were subsequently converted to Fisher z-scores for further analysis. The atlas-defined WM tract ROIs were defined by 48 regions of interest (ROIs) from the JHU ICBM-DTI-81 WM atlas (Mori et al., 2008). The GM parcels comprised 90 ROIs derived from the Automated Anatomical Labeling (AAL) atlas template (Tzourio-Mazoyer et al., 2002). In addition, these 90 GM brain regions were further divided into 7 GM functional sub-networks (Yeo et al., 2011), including the Attention Network (AN), Basal Ganglia Network (BGN), Default Mode Network (DMN), Frontoparietal Network (FN), Limbic Network (LN), Sensorimotor Network (SN), and Visual Network (VN). All atlas-defined WM and GM regions(defined in MNI space) were intersected with the corresponding normalized tissue masks to constrain fMRI signal extraction within tissue-specific regions.. These masks were generated by thresholding the TPMs at 0.8, applying a stricter criterion than the initial segmentation to prevent signal contamination between WM and adjacent GM.

With these masks, the functional networks were constructed in two forms. The intrinsic WM-WM connectivity was represented by a symmetric 48 × 48 adjacency matrix. For the WM-GM network, to satisfy the input requirements of NBS for symmetric matrices, following Xu et al. (2024), the asymmetric WM-GM connection (size=48×90) was constructed as a virtual symmetric matrix (size=138×138) by combining both WM nodes (n=48) and GM nodes (n=90) as virtual nodes.

To reduce potential biases from multi-site acquisition effects, a ComBat harmonization procedure (Johnson et al., 2007) was implemented using the neuroCombat package, modeling acquisition site as the batch effect, while age, sex, head motion, and diagnostic group (ASD/TC) were included as biological covariates. Briefly, the ComBat model describes the rsFC values in the form *y_ijv_* = *α_v_* + *X_ij_*/*β_v_* + *y_iv_* + *o_iv_ E_ijv_* where *y_ijv_* corresponds to the rsFC measurement for the *i* acquisition site, *j* subject, and *v* WM-GM or WM-WM connection. The term α*_v_* denotes the population-level mean connectivity for connection *v*, while *X_ij_* constitutes the covariate design matrix encoding biological confounders. The term *y_iv_* and *δ_iv_* capture site-specific additive bias and scaling factors respectively, with *ε_ijv_* representing stochastic noise components. The harmonized rsFC values were then computed as 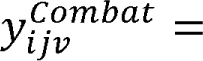 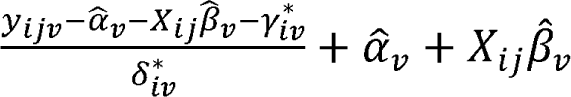 where 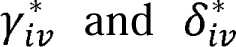 represent stabilized estimates of site effects through posterior maximization. Crucially, incorporating age, sex, and head motion as covariates preserved biologically meaningful rsFC variations associated with these factors while removing non-biological site effects.

### 2.4 Statistical Analysis

#### 2.4.1 Network-Based Statistic for Group Comparison

To examine between-group differences (ASD group vs. TC group) in WM-GM FC and WM-WM FC, this study employed the Network-Based Statistic (NBS) method for group comparisons at a significance level of P<0.05. NBS is a validated statistical approach for detecting differences in network data between two or more groups, particularly prevalent in neuroimaging research and brain network analysis (Zalesky et al., 2010).

For WM-WM network, analysis was conducted on the 48×48 WM-WM symmetric adjacency matrix. First, a two-sample t-test was applied to each pairwise FC using an initial t-value threshold of 3.1, a value that approximates an uncorrected p < 0.001 and was used to generate the initial suprathreshold components. Then, the Family-Wise Error Rate (FWER)-corrected threshold of P were calculated for each component using a permutation test (5000 permutations; significance level = 0.05). Using this NBS method, we identified WM-WM pairs with significantly disrupted FC values in the test group relative to the control group.

For the WM-GM network, analysis was conducted on the 138×138 virtual symmetric matrix. The WM-WM (upper left 48×48) and GM-GM (lower right 90×90) regions were zeroed out by masking to avoid interference within the WM or within the GM. The subsequent analysis steps were consistent with WM-WM.

To further summarize these changes at the network level, we calculate the ratio of the number of anomalous connection pairs within a particular network to the total number of nodes in that network. This normalization enables an unbiased comparison of anomaly profiles across networks of varying sizes.

#### 2.4.2 Correlation Analysis with the Severity of Clinical Symptoms

To assess the clinical relevance of the identified network alterations, we performed Pearson correlation analyses between the connectivity strength of each aberrant edge from the NBS results and the SRS total scores. Prior to correlation, the effects of potential confounders, including age, sex, and mean FD, were regressed out from each connectivity value using a linear model. The resulting P-values were corrected for multiple comparisons across all tested connections using the False Discovery Rate (FDR) method (q < 0.05)

### 2.5 Machine Learning

To evaluate the classification potential of abnormal brain connections identified via the NBS method, we constructed a CatBoost classifier using WM-WM, WM-GM, and their combined connections as input features. CatBoost was chosen because it performs well on datasets where the number of input features is relatively large compared to the number of samples (i.e., a moderately high-dimensional brain connectivity scenario), shows robustness to multicollinearity, and generally offers better generalization compared with conventional tree-based models (Prokhorenkova et al., 2018). In particular, similar studies in connectomics have shown successful application of CatBoost on brain network data (Zhang et al., 2023), and reviews of machine learning in macroscale connectomics report that the feature-space in such data tends to be high-dimensional and complex (Anbarasi et al., 2024).

Feature importance was first computed using a random forest algorithm within each training fold of the five-fold cross-validation, and features were ranked accordingly. This ensures that feature ranking is based solely on training data and avoids data leakage. We used the random forest algorithm instead of CatBoost’s built-in importance calculation because it provides a model-agnostic and interpretable measure of variable relevance, independent of the boosting process in CatBoost, thereby reducing potential bias in feature selection. Previous studies have shown that random forest–based importance measures (e.g., mean decrease in impurity) are stable and highly interpretable across datasets (Agarwal et al., 2023). Furthermore, different algorithms often produce inconsistent importance rankings due to model-specific mechanisms, and using a model-agnostic approach helps to ensure unbiased feature selection (“Comparison of feature importance measures as explanations for classification models,” Herman et al., 2021). In addition, recent work has demonstrated that model-agnostic ranking strategies improve generalization in high-dimensional small-sample conditions (Li et al., 2024). To minimize variance due to data partitioning, the entire pipeline—including feature selection, tuning, and evaluation—was repeated ten times with different random seeds. Performance metrics (accuracy, AUC, sensitivity, specificity) were averaged across repetitions to provide robust assessment.

## 3. Result

### 3.1 Harmonization

Possible site-effects on WM-WM rsFC were effectively corrected by the harmonization process which mitigated mean and variance differences across 13 sites, as shown in the comparison of rsFC at acquisition sites before and after harmonization (Fig. 1A&B). Similarly, site-related variability in WM-GM rsFC was also successfully mitigated (Fig. 1C & D). Notably, the median connectivity strength at each site remained relatively stable following correction: WM-WM rsFC centered around 0.5, while WM-GM rsFC consistently ranged between 0.3 and 0.4. This indicates that the harmonization process effectively reduced inter-site variability without substantially altering the central tendency of the functional connectivity data.

**Fig. 1.**
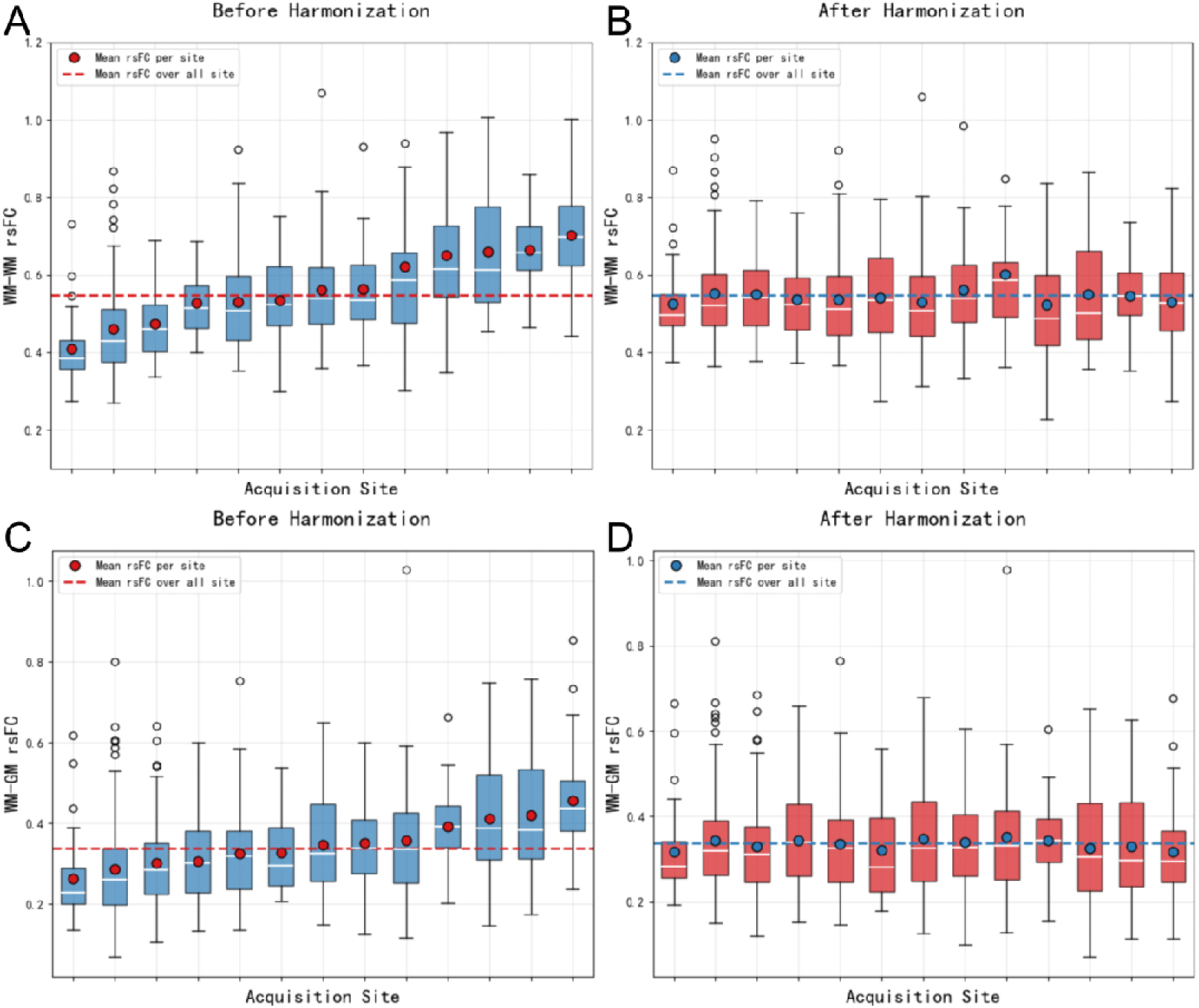
Comparison of summary statistics (including median, 0.25 and 0.75 quartiles, non-outlier minima and maxima, and outliers) for rsFC at 13 collection points before and after harmonization. Colored dots indicate the mean rsFC within a collection point. The colored dashed line is the reference line for the rsFC average across all collection points.

### 3.2 Alteration of WM–WM FC in ASD

NBS identified 116 significant WM-WM connectivity pairs in ASD patients compared to the TC group (FWER-corrected P<0.001). Notably, all significantly abnormal connections were enhanced. As shown in Fig. 2 (A & C), these anomalous WM-WM pairs involved 25 out of 48 WMs (52.1%), and the frequency of connectivity anomalies was significantly higher in some WM tract ROIs than in other regions. The number of abnormal connections in Tapetum R (T.R; 20 times), Posterior corona radiata L (PCR.L ; 18 times) and Anterior corona radiata R (ACR.R ; 17 times) were the most prominent, and these regions may serve as key nodes for information transmission in autistic patients. In addition, we identified the three most connected pairs among all the significant connections: T.R and Anterior corona radiata L (ACR.L; t=5.893), PCR.L and Fornix (FX; t=5.793), and ACR.R and T.R (t=5.715).

**Fig. 2.**
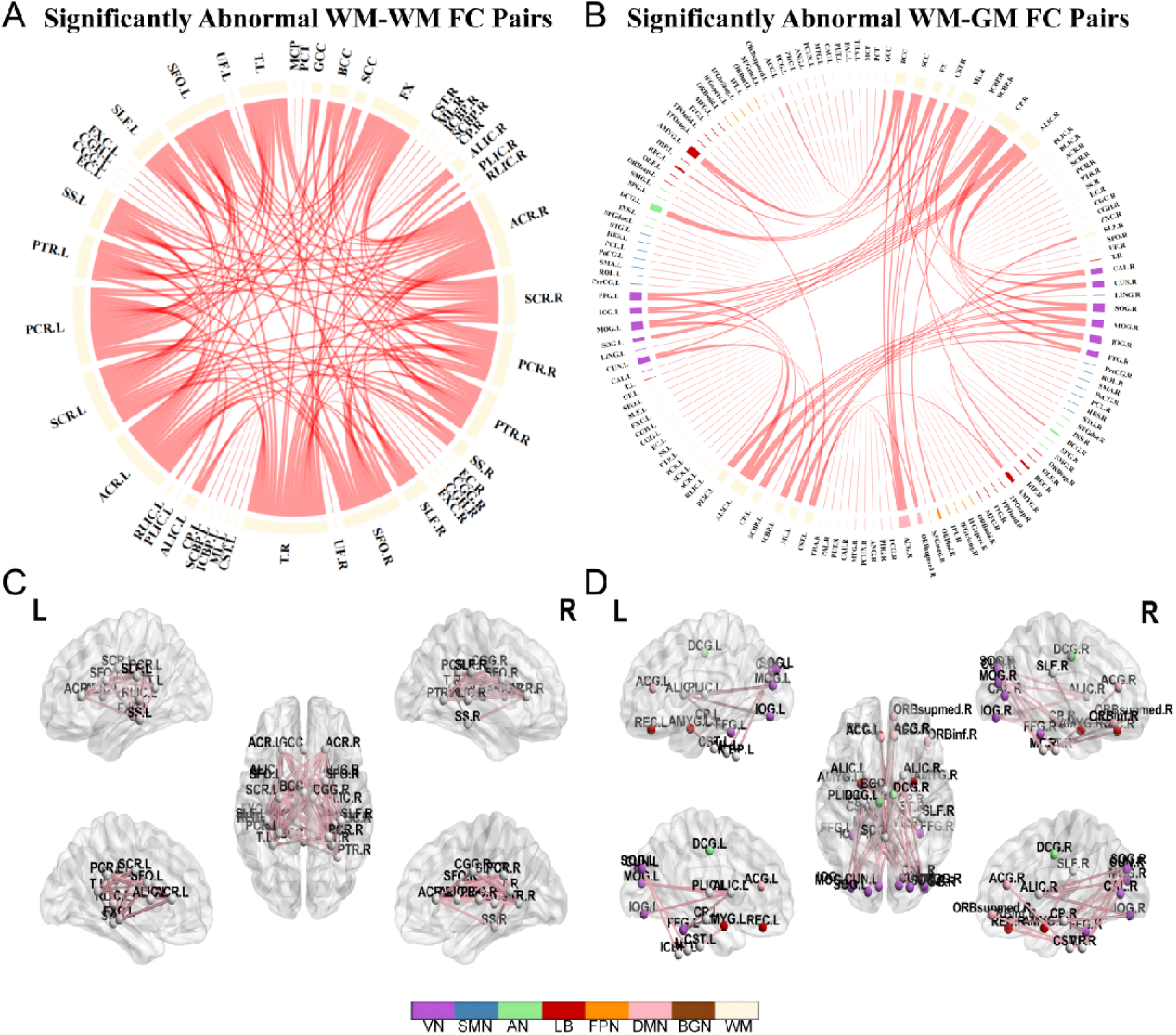
Significantly abnormal linkage pairs identified by NBS. (A) Significantly abnormal WM-WM linkage pairs. The thickness of the line represents the magnitude of the effect, and the red line indicates a positive correlation. (B) Significantly anomalous WM-GM linkage pairs. The thickness of the line reflects the effect size, and the red line represents enhancement. (C) Identification of 25 nodes within the group-averaged WM-WM network, underscoring their critical roles in the brain’s functional connectivity architecture. (D) Identification of 36 nodes within the group-averaged WM-GM network.

#### 3.2.1 Alteration of WM–GM FC in ASD

NBS identified 58 significant WM-GM connectivity pairs (FWER-corrected P=0.018) in agreement with WM-WM, all 58 of which are enhanced. As illustrated in Fig.2(B & D), these abnormal WM-GM pairs involved 21 of 90 GM nodes (36.2%) and 15 of 48 WM tract ROIs (31.3%). According to the classification of GM, it can be corresponded to four Yeo network classification functional networks (VN, LN, AN, DMN), while the percentage of abnormal connection pairs in VN is higher than other functional networks (78.6%). In addition, we found that ASD exhibited significantly elevated FC between Cingulum Ant R (ACG.R) and Splenium of corpus callosum (SCC) compared to the normal group (t = 3.9752).

### 3.3 Investigating Symptom Severity Correlates in Clinical Presentations

To explore the neural correlates of clinical symptom severity, we perform a correlation analysis between previously identified white matter connectivity abnormalities and SRS total scores. This investigation was conducted at two resolutions. First, at the whole-brain level, we assessed the overall relationship. The mean strength of WM-WM functional connections constituting the entire previously identified abnormal network showed a significant negative correlation with SRS total scores (r = -0.22, p < 0.001, N = 456; Fig.3A). This global finding indicates that, overall, reduced strength within this abnormal network is associated with elevated autism symptom severity. To further delineate which specific connections within this abnormal network contribute most to this overall association, we proceeded with an edge-by-edge analysis. For this, we took the 116 connections previously identified as exhibiting significant abnormalities and correlated each one individually with SRS total scores. After FDR correction of these 116 individual correlation analyses, 85 WM-WM connections maintained a significant association with SRS total scores. Among these 85 connections, the most pronounced effects emerged in limbic-visual pathways. Specifically, the strongest negative correlations were observed between the left anterior corona radiata (ACR.L) and right tapetum (Tapetum.R) (r = -0.223, p = 0.0001), followed by left superior fronto-occipital fasciculus (SFO.L) to Tapetum.R connectivity (r = -0.221, p = 0.0001). Additional significant associations included ACR.R to Tapetum.R (r = -0.197, p = 0.0008), right superior fronto-occipital fasciculus (SFO.R) to Tapetum.R tracts (r=-0.195, p=0.0008), and left superior corona radiata (SCR.L) to Tapetum.R connectivity (r=-0.184, p=0.0018).

**Fig. 3.**
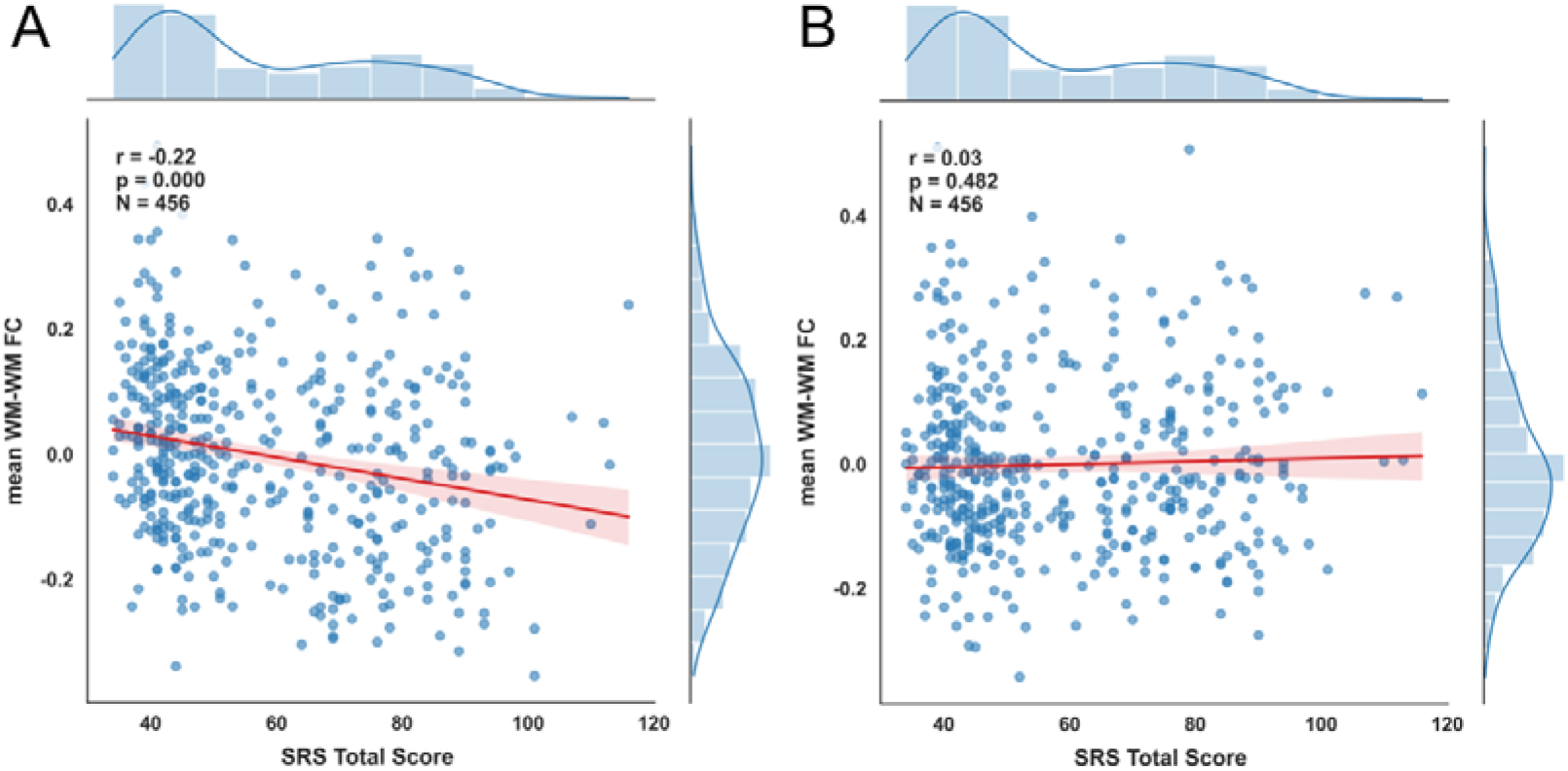
Correlation analysis with the severity of clinical symptoms (A) Colleration between Mean WM-GM FC and SRS Total Score. (B) Colleration between Mean WM-WM FC and SRS Total Score.

**Fig. 4.**
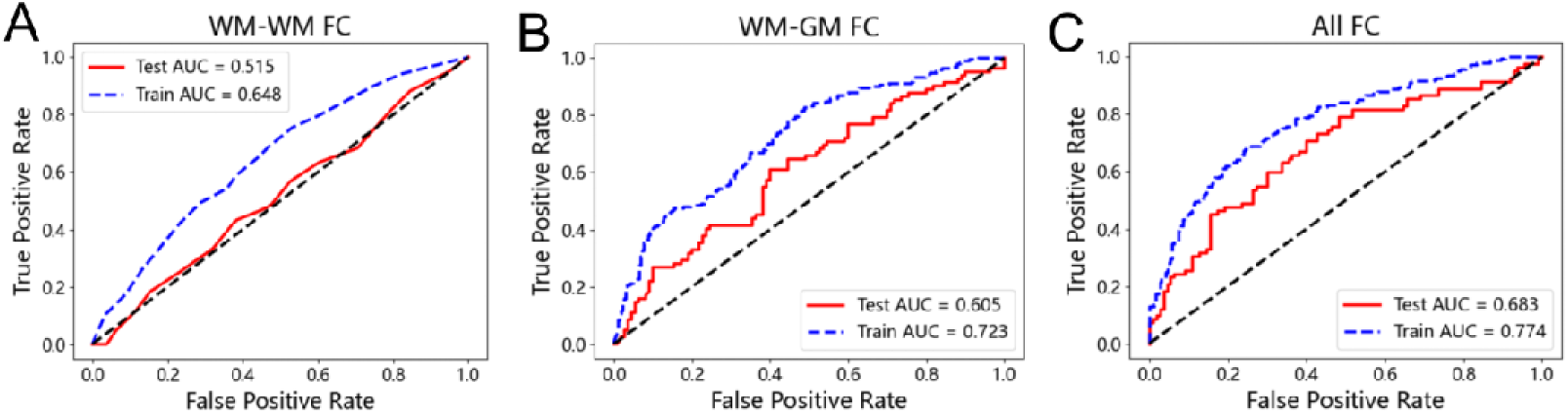
Receiver operating characteristic (ROC) curves illustrating the classification performance of models based on different functional connectivity (FC) features. (A) Model trained on white matter–white matter (WM-WM) FC features yielded an area under the curve (AUC) of 0.515 on the test set and 0.648 on the training set. (B) Model based on white matter–gray matter (WM-GM) FC features achieved a test AUC of 0.605 and a training AUC of 0.723. (C) Combined model incorporating both WM-WM and WM-GM FC features attained the highest performance, with a test AUC of 0.683 and a training AUC of 0.774.

In contrast, no significant correlations emerged between WM-GM connectivity and SRS total scores. Both edge-by-edgevoxel-wise analyses of individual WM-GM connections and aggregate mean connectivity values (r = 0.03, p = 0.482, N = 456; Fig.3B) revealed null associations with SRS total scores.

### 3.4 Evaluation of the Ability to Categorize Anomalous Connections

The classification results revealed that combining abnormal WM-WM and WM-GM connections led to superior performance compared to using either connection type alone. The combined model achieved an average accuracy of 0.659 ± 0.023, AUC of 0.669 ± 0.040, sensitivity of 0.316 ± 0.045, and specificity of 0.912 ± 0.034 across ten repetitions. In contrast, the WM-WM model yielded lower accuracy (0.558 ± 0.016) and AUC (0.547 ± 0.054), with extremely low sensitivity (0.022 ± 0.040) despite high specificity (0.968 ± 0.054). The WM-GM model showed similar patterns, with accuracy of 0.573 ± 0.011, AUC of 0.589 ± 0.043, sensitivity of 0.024 ± 0.026, and specificity of 0.979 ± 0.022.

## 4. Discussion

This study investigated the functional architecture of WM networks in ASD, focusing on abnormalities within WM-WM and WM-GM connectivity and their clinical relevance. Our analysis of rs-fMRI data from the ABIDE-II cohort revealed three primary findings: First, utilizing Network-Based Statistics, we identified widespread and significant enhancements in FC across numerous WM-WM (116 pairs) and WM-GM (58 pairs) connections in individuals with ASD compared to typical controls. Second, these aberrant intra-WM (WM-WM) connections were found to be clinically significant, as their increased strength showed a robust negative correlation with autism symptom severity, measured by the SRS; this association was not observed for WM-GM connectivity. Third, a machine learning model (CatBoost) integrating features from both WM-WM and WM-GM networks demonstrated moderate diagnostic accuracy (AUC = 0.669 ± 0.040) in distinguishing ASD individuals from controls, outperforming classifiers based on single connectivity types. These results collectively provide novel evidence for specific dysregulation within WM functional networks as a significant facet of ASD neuropathology, directly linking these alterations to social-communicative deficits and highlighting their potential as integrated biomarkers.

### 4.1 Widely enhanced white matter connectivity and its key nodes

We identified 116 significantly enhanced WM-WM connections in ASD. These findings, combined with Boucher et al.’s (2007) observations of abnormal WM connectivity during early neurodevelopmental windows, suggests that abnormalities in the white matter network may be an earlier and more prominent pathologic feature of ASD. This finding supports Arunachalam Chandran et al.’s (2021) suggestion that “white matter abnormalities may be one of the major etiologic factors and influences in ASD”.

Notably, 52.1% of affected WM nodes showed exceptionally strong connections (t-values >5.7), indicating that traditionally dismissed “noise” in WM signals actually contains critical pathological information (Ji et al., 2023). In particular, three hub nodes (T.R, PCR.L, ACR.R) formed a cross-modal network through three high-significance connections: ACR.L-T.R (t=5.893), PCR.L-FX (t=5.793), and ACR.R-T.R (t=5.715). This distinctive structural pattern provides new mechanistic insights into ASD core symptoms.

Specifically, extensive connectivity enhancement of T.R nodes may disrupt the function of the thalamus in information processing, triggering abnormal integration of sensory information, (Sherman et al., 1998; Guillery et al., 2002), which is clinically manifested as hypersensitivity or unresponsiveness to external stimuli. Whereas ACR.R and PCR.L act as projection fibers connecting the cerebral cortex to the thalamus, brainstem, and spinal cord, which are responsible for transmitting motor and sensory signals. Their damage may lead to irreversible neurological deficits (Yakar, F et al., 2018). These findings systematically reveal the central role of white matter networks in ASD, making white matter network parameters more clinically promising biomarkers.

### 4.2 Visual Network Hyperconnectivity and Synergistic Anomalies

Our results demonstrate that brain network disruption in ASD is not uniform across functional systems. Specifically, abnormal WM-GM connectivity in ASD patients was mainly concentrated in the visual network, and all showed enhanced functional connectivity. This finding is highly consistent with the functional connectivity study of Keehn et al. (2013) for the ASD visual search task, but differs from the phenomenon of weakened connectivity that is common to defective domains in previous studies. This paradox coincides with the implicit inference in the “low connectivity theory” proposed by Just et al.’s (2023). While the theory primarily explains ASD’s functional deficits in complex cognitive tasks, its core mechanisms are equally applicable to the domain of dominance: the visual network relies on WM-GM’s unusually enhanced local connectivity for efficient detail processing by reducing the need for cross-network integration.

In addition, we identified a significant enhancement of ACG.R and SCC connectivity. As a key node of the default mode network (Uddin et al., 2009), ACG.R has been shown to be associated with self-referential processes and social cognition (D’Argembeau et al., 2005; Iacoboni et al., 2004;)----which are precisely the areas typically impaired by ASD. Combined with the key role of the SCC in visuospatial information transfer (Knyazeva et al., 2008), the enhancement of this connection may reflect an unusual synergistic mechanism othe enhancement of this connection may reflect an unusual synergistic mechanism of spontaneous thinking and visuospatial cognitive functioning.

### 4.3 Correlation analysis with the severity of clinical symptoms

Our investigation demonstrates a robust inverse association between WM inter-network functional connectivity and socio-communicative symptom severity in ASD, as quantified by SRS. Notably, the most pronounced correlations emerged in 85 anatomically distinct pathways. The most relevant are the ACR.L and Tapetum.R connections. These observations extend previous neuroanatomical findings by establishing functional correlates of WM integrity deficits in core ASD symptomatology.

The neurobiological significance of these findings becomes apparent when considering the functional roles of the implicated tracts. The anterior corona radiata, as a critical component of cortical-subcortical regulatory circuits, mediates emotional modulation and cognitive control through its widespread projections to prefrontal and limbic regions (Sanjuan et al., 2013; Choi et al., 2015). Compromised connectivity in this pathway could impair hierarchical information processing, potentially manifesting as disrupted sensory gating and emotional dysregulation - hallmarks of ASD psychopathology. Similarly, the tapetum’s essential role in interhemispheric integration of temporo-parietal networks (Hofer & Frahm, 2006; Park et al., 2008) suggests that its reduced connectivity may contribute to impaired cross-modal synthesis of social cues and linguistic information, providing a neural substrate for characteristic communication deficits in ASD. These findings corroborate previous reports of WM microstructural abnormalities in ASD, particularly in tracts linking socio-emotional and sensory-processing regions (Keller et al., 2007; Shukla et al., 2011).

The absence of significant correlations between WM-GM connectivity and SRS total scores stands in contrast to the significant associations observed within the WM-WM network. This discrepancy may be attributed to several methodological and neurobiological factors. Firstly, from a methodological perspective, WM-GM connectivity, by definition, represents a direct interface between two functionally distinct tissue types. The BOLD signal in this transitional zone might be more susceptible to partial volume effects and other sources of physiological noise, which could obscure subtle, behaviorally-relevant functional relationships (Peer et al., 2017). More importantly, from a neurobiological standpoint, our findings suggest that the social deficits measured by the SRS in ASD may be more closely linked to the efficiency and integrity of long-range communication within the distributed white matter infrastructure itself, rather than to the specific functional coupling at the WM-GM junctions. Converging evidence indicates that WM-WM connections reflect the synchronized electrophysiological activity of macro-scale fiber tracts (Huang et al., 2023), which could underpin broader network dynamics relevant to complex social behavior (Zu et al., 2024). The lack of association with WM-GM connectivity does not diminish the role of gray matter but indicates that the clinical symptoms captured by the SRS in our cohort are more robustly reflected in the intrinsic functional organization of the white matter networks we identified.

Although NBS analysis revealed significantly enhanced WM connectivity in ASD relative to controls, correlation analyses within the ASD cohort demonstrated that individuals with stronger connectivity in these same edges exhibited lower SRS scores. This paradox can be reconciled by considering compensatory mechanisms (Uddin et al., 2013), whereby certain ASD individuals may increase connectivity in specific pathways to offset deficits, resulting in milder behavioral symptoms (Plitt et al., 2015). Moreover, these connections may be primarily strengthened in mild ASD subgroups, creating a shift in group means while driving an inverse relationship within the ASD group. Importantly, Nair et al. (2015) identified similar patterns in thalamocortical pathways, where hyperconnectivity predicted better social communication, supporting the notion that not all connectivity enhancements are pathological. These insights illustrate the importance of integrating group-level contrasts and individual-level correlations to fully understand ASD neurobiology.

### 4.4 Machine learning

This study systematically investigated the role of white matter–white matter (WM-WM) and white matter–gray matter (WM-GM) functional connectivity features derived from fMRI in the classification of autism spectrum disorder (ASD). For the first time, abnormal connections from both types were combined for model construction. The results demonstrated that the combined model outperformed either WM-WM or WM-GM alone in terms of classification accuracy and AUC, suggesting that these two types of abnormal connectivity may provide complementary information and serve as potential biomarkers of ASD.

Previous studies, as shown in Table 2, have primarily focused on using resting-state FC between GM regions as model features. (Heinsfeld et al., 2018; Abraham et al., 2017). For example, Heinsfeld et al. achieved over 70% classification accuracy using an autoencoder-based model on the entire ABIDE dataset (n = 1035), while Abraham et al. (2017) reported approximately 67% accuracy using PCA and SVM. In contrast, the present study introduced a novel feature construction approach based on fMRI-derived white matter signals and incorporated abnormal WM-WM and WM-GM connections in a unified framework. Despite a smaller sample size (n = 640) and stringent feature selection, the combined model achieved an average accuracy of 0.659 ± 0.023 and an AUC of 0.669 ± 0.040, highlighting the potential utility of white matter functional connectivity in ASD classification.Although deep learning models trained on large-scale datasets have reported higher accuracy, they are often prone to overfitting (Arbabshirani et al., 2017) and suffer from poor interpretability, which limits their clinical utility. In contrast, our study adopted a lightweight classification framework with controlled feature dimensions and rigorous cross-validation, resulting in a balanced outcome in terms of accuracy, stability, and biological interpretability.

**Table 2.**
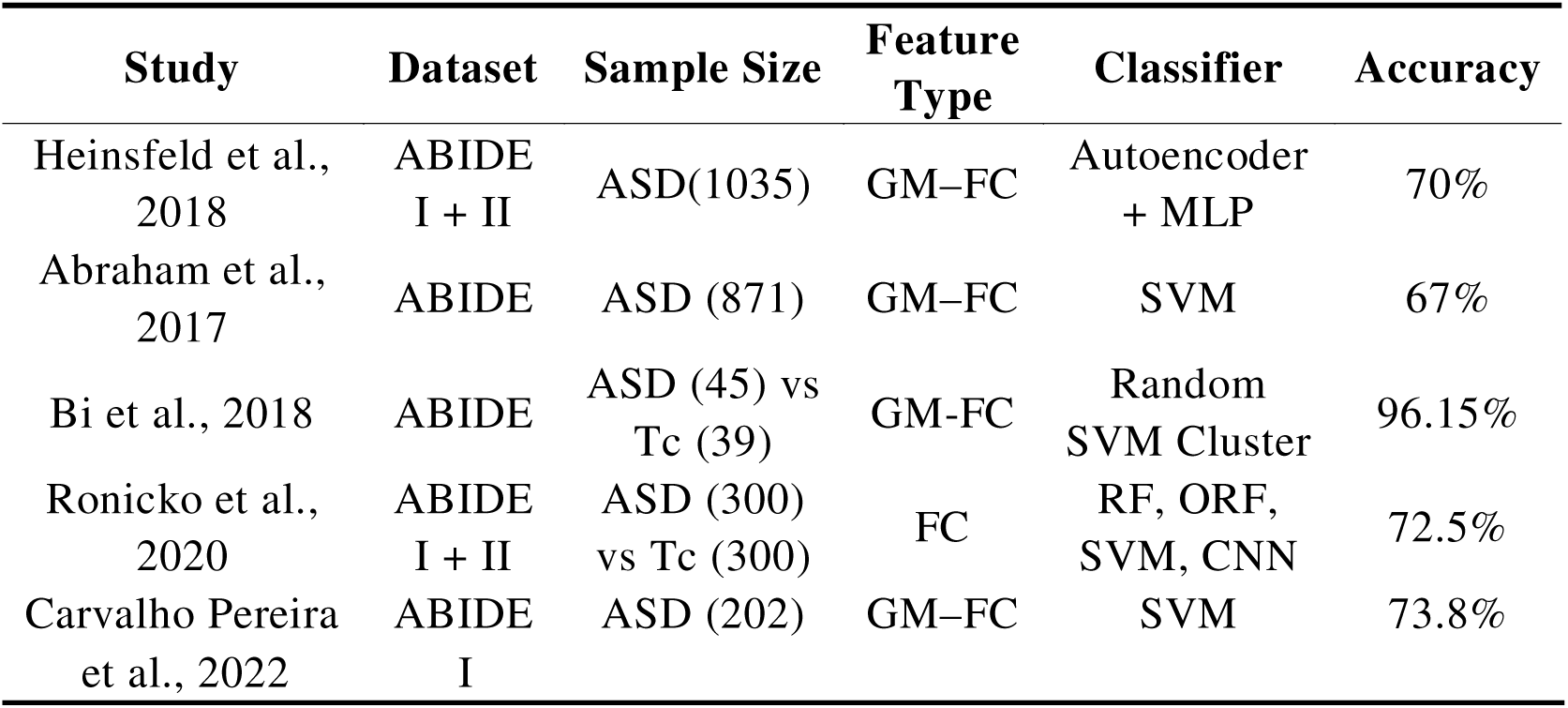
Methodological comparison of representative machine-learning studies for ASD classification using rs-fMRI.

Compared with other studies using similar sample sizes, our results are also competitive. However, direct numerical comparison should be treated with caution because prior ASD studies differ substantially in sample characteristics, feature modalities, FC estimation strategies, and classifier designs. For instance, Plitt et al. (2015) used GM-based FC and traditional SVM classifiers on a combined in-house and ABIDE cohort with different sample compositions, reporting accuracies ranging from 60% to above 70% rather than a single fixed value. Likewise, Riaz et al. (2020) performed multimodal fusion (fMRI + sMRI) using deep learning architectures, which fundamentally differs from our single-modality WM-based approach. These discrepancies illustrate that distinct methodological choices—rather than model superiority—largely account for performance variation across studies.

A broader view of machine-learning studies further supports this point. For example, Bi et al. (2018) applied a Random SVM Cluster ensemble on a much smaller subset of ABIDE (n = 84 after motion exclusion), achieving up to 96.15% accuracy, demonstrating how small-sample ensemble methods can produce inflated performance under highly optimized feature subsets. Ronicko et al., 2020 compared multiple FC estimation methods (GLASSO, MDMC, Pearson correlation) and classifiers (RF, ORF, SVM, CNN), and achieved single-trial accuracy of 72.5% and AUC around 0.73, showing that choices in FC computation and model architecture significantly affect results. Carvalho Pereira et al. (2022) focused on ASD severity classification (a different task from ASD–control discrimination), achieving 73.8% accuracy using GM-FC features, highlighting that even the prediction objective varies across studies.

Within this heterogeneous methodological landscape, the present study provides a complementary contribution by focusing on white matter–derived functional connectivity—an underexplored modality in ASD machine-learning research. By integrating NBS-identified abnormal WM-WM and WM-GM connections with a controlled feature selection pipeline and CatBoost modeling, our results suggest that white matter functional connectivity encodes discriminative information that is not captured by gray matter features alone. Rather than emphasizing absolute performance, our findings reinforce that ASD classification is jointly shaped by sample heterogeneity, feature construction, FC estimation, and classifier choice, and demonstrate the potential value of incorporating white-matter FC into future ASD biomarker investigations.

### 4.5 Limitation

Although advancements have been achieved in this study, several limitations and potential avenues for refinement warrant consideration. First, the cross-sectional design based on data snapshots inherently restricts our capacity to elucidate dynamic alterations in WM and GM functional networks among individuals with ASD. Second, the exclusive reliance on 640 samples from the ABIDE II public database introduces constraints regarding sample size adequacy and methodological rigor. The absence of an independent internal validation cohort and supplementary verification using institutional data may compromise the model’s generalizability and clinical translatability. Future investigations should prioritize sample size expansion and incorporation of multimodal neuroimaging data to enhance model robustness. Third, despite sex-matching between ASD and TC groups, the pronounced male predominance in both cohorts necessitates future studies to ensure greater inclusion of female participants. Finally, given the substantial heterogeneity inherent in ASD manifestations, subsequent research should emphasize individual variability characterization to facilitate precision therapeutic interventions.

## Conclusion

In summary, this study constructed WM-related functional networks in ASD, which are involved in sensory and socio-cognitive processing. Crucially, aberrant functional connectivity specifically within the WM networks exhibits a robust negative correlation with ASD symptom severity, whereas WM-GM functional connectivity demonstrates a distinct clinical dissociation. This dissociation underscores that intrinsic dysfunction within the WM networks themselves represents a distinct pathological mechanism contributing to the social and communication difficulties characteristic of ASD. Furthermore, the integrated analysis of connectivity features from both WM and GM compartments demonstrates significant diagnostic potential, surpassing approaches focusing on single networks and reinforcing the clinical utility of mapping whole-brain functional connectivity. Collectively, these findings establish intrinsic white matter functional networks as a critical neural substrate in ASD pathophysiology, linking WM structural alterations to core clinical manifestations and providing novel targets for mechanistic exploration.

## Ethical Statement

All procedures performed in studies involving human participants were in accordance with the ethical standards of the institutional and/or national research committee and with the 1964 Helsinki declaration and its later amendments or comparable ethical standards. This article does not contain any studies with animals performed by any of the authors. Informed consent was obtained from all individual participants included in the study.

## Authors Contribution

Ze-Qiang Linli conceived and designed the overall study. Si-Jing Wu, Miao-Ting Huang, and Dan-Wei Huang oversaw data collection and verified the analytical methods. Si-Jing Wu, Ze-Qiang Linli, Shui-Xia Guo performed data preprocessing, constructed the networks, and carried out the statistical analyses. Si-Jing Wu, Miao-Ting Huang, and Dan-Wei Huang contributed to the methodology design and drafted large parts of the manuscript. Ze-Qiang Linli provided critical revisions, shaped the discussion of results, and coordinated manuscript editing. All authors discussed the results, commented on the manuscript at all stages, and approved the final version for publication.

## Conflict of Interest Statement

The authors declare that they have no conflict of interest.

## Financial support

Ze-Qiang Linli is supported by the Humanities and Social Sciences Youth Foundation of Ministry of Education of China (24YJCZH164), and the Guangdong Province General University Characteristic Innovation projects (2024KTSCX178).

## Data Availability

https://fcon_1000.projects.nitrc.org/indi/abide/

## Notes

### Competing Interest Statement

The authors have declared no competing interest.

### Funding Statement

This work was supported by the Humanities and Social Sciences Youth Foundation of Ministry of Education of China (No. 24YJCZH164 to Z.L.); the National Training Program of Innovation and Entrepreneurship for Undergraduates (No. 202511846014); the Guangdong Province General University Characteristic Innovation projects (No. 2024KTSCX178 to Z.L.); and the Provincial Training Program of Innovation and Entrepreneurship for Undergraduates (No. 202511846061).

### Author Declarations

https://fcon_1000.projects.nitrc.org/indi/abide/

